# COVID-19 in a Rural Health System in New York – Case Series and an Approach to Management

**DOI:** 10.1101/2020.10.15.20213348

**Authors:** Eyal Kedar, Regina Scott, Daniel Soule, Carly Lovelett, Kyle Tower, Kylie Broughal, Daniel Jaremczuk, Sara Mohaddes, Imre Rainey-Spence, Timothy Atkinson

**Affiliations:** Division of Rheumatology, Department of Medicine, St. Lawrence Health System, Potsdam, NY; Department of Pharmacy, St. Lawrence Health System, Potsdam, NY; Division of Infectious Disease, Department of Medicine, St. Lawrence Health System, Potsdam, NY; Department of Clinical and Rural Health Research, St. Lawrence Health System, Potsdam, NY; Division of Hospital Medicine, Department of Medicine, St. Lawrence Health System, Potsdam, NY

## Abstract

**Background:** Many rural hospitals and health systems in the U.S. lack sufficient resources to treat COVID-19. We developed a system for managing inpatient COVID-19 hospital admissions in St. Lawrence County, an underserved rural county which is the largest county in New York State.

**Methods:** We used a hub and spoke system to route COVID-19 patients in the St. Lawrence Health System to its flagship hospital. We assembled a small clinical team to manage admitted COVID-19 patients and to stay abreast of a quickly changing body of literature and standard of care. We subsequently completed a review of clinical data for patients who were treated by our inpatient COVID-19 treatment team between March 20 and May 22, 2020.

**Results:** Twenty COVID-19 patients were identified. Sixteen patients (80%) met NIH criteria for severe or critical disease. One patient died. No patients were transferred to other hospitals.

**Conclusions:** During the first two months of the pandemic, we were able to manage hospitalized COVID-19 patients in our rural community. Development of similar treatment models in other rural areas should be considered.

## INTRODUCTION

As of October 2nd, 2020, there have been almost 35 million cases and over one million deaths globally from Severe Acute Respiratory Syndrome Coronavirus-2 (SARS-CoV-2). Of these, over seven million cases are in the U.S.. Rural areas of the U.S. ^1^ have higher rates of older patients and patients with multiple medical comorbitidies^2-3^. This, coupled with a severe and chronic shortage of medical subspecialists^3^, relative shortages of both regular hospital beds and ICU beds^4^ and the accelerating rate of financial failure of many rural hospitals^5^ has left a large part of the rural U.S. without sufficient hospital bed capacity, personal protective equipment (PPE), medications or access to subspecialists to treat some of the sickest patients with COVID-19.

Rural hospitals also face another underappreciated but very significant challenge in treating COVID-19 patients. In most rural hospitals in the U.S., there is a lack of the experience and/or infrastructure necessary to participate in clinical trials^6-7^. This leaves many rural physicians with the choice of either treating their COVID-19 patients with off label medications which, according to various society guidelines, should be used only in the context of clinical trials^8-9^, or of not treating their patients at all.

This report aims to describe the St. Lawrence Health System’s (SLHS) model for inpatient COVID-19 care delivery, including the assembly of a small team of specialists with broad comfort within their respective disciplines and a hub and spoke model for transferring patients within the SLHS to its flagship hospital, Canton Potsdam Hospital (CPH). In the context of this care delivery system, it further aims to describe the demographic characteristics and clinical course of the 20 COVID-19 patients admitted to CPH over the first two months of the COVID-19 pandemic. The final aim of the report is to discuss strategies for implementing similar care delivery strategies in other rural areas both during this and future pandemics.

## METHODS

As part of its COVID-19 triage plan, the SLHS uses a hub and spoke model in which all suspected COVID-19 cases are transferred to its flagship hospital, CPH. We assembled a clinical team consisting of an infectious diseases physician and advanced practice provider, a rheumatologist with experience in managing interstitial lung disease, a hospitalist team with experience in critical care medicine and an antimicrobial stewardship pharmacist with experience in general inpatient pharmacy. Since its inception, this team has met daily to conduct multidisciplinary team rounds for all hospitalized COVID-19 patients at CPH. As part of these rounds, the team also discusses COVID-19 research updates, epidemiologic data updates for our region and COVID-19 medication inventory updates for our pharmacy. The date of data censoring for this study coincides with the date on which we began our first inpatient COVID-19 clinical trial at CPH, and to this end our daily rounds also include a discussion of both new and existing COVID-19 trial opportunities through our health system’s clinical research department.

This study was conducted at CPH during the initial phase of COVID-19 infection in St. Lawrence County (SLC) between 3/20/2020 and 5/22/2020. We included all consecutive adult (>age 18) patients with confirmed and active COVID-19 infection who were admitted to CPH over the dates of the case series. A confirmed active COVID-19 case was defined as having a positive RT-PCR assay on a specimen obtained via nasopharyngeal swab and sent to a reference lab for testing for SARS-CoV-2 RNA. Patients with positive serologic but negative RT-PCR testing at the time of admission were excluded. Twenty adults were identified, none of whom were pregnant or prisoners. The study was approved by the SLHS institutional review board. A waiver of consent was obtained for patients lost to follow-up. All other patients provided written authorization to use and share health information. Clinical outcomes were monitored until the final date of follow-up on 5/22/2020.

Data were collected from CPH’s electronic medical record system (Meditech and ECW). Data collected on admission included demographic information, baseline comorbidities, symptoms prior to admission and pertinent travel and COVID-19 exposure histories. Data collected over the course of the hospital stay included vital signs, lab results, chest imaging results, echocardiogram and electrocardiogram results, inpatient medications and a daily NIH ordinal scale score for all patients.

Measured clinical outcomes included length of stay and death rate. Clinical measures included severity of illness, supplemental oxygen requirements, need for invasive mechanical ventilation, use of vasopressors, acute kidney injury (defined as an increase in serum creatinine of ≥0.3 mg/dL within 48 hours or ≥50% within 7 days or urine output of <0.5 mL/kg/hour for >6 hours^10^) and transaminitis.

Statistical analysis was performed using R software (version 3.3.2). Results were reported as means and standard deviations, medians and interquartile ranges or, in the case of categorical variables, counts and percentages. There were no missing data for which imputation was necessary.

## RESULTS

### Demographics and baseline clinical characteristics

The demographic and baseline clinical characteristics of the 20 COVID-19 positive patients admitted to CPH between March 20 and May 22, 2020 are shown in Table 1. The median age of the patients was 63 (IQR 51-74) with 12 (60%) females. Ten (50%) patients were obese, four patients (20%) had cardiovascular disease, two (10%) had diabetes mellitus, four (20%) had pulmonary disease (COPD or asthma) and four (20%) had obstructive sleep apnea. Six (30%) patients were current or former smokers. The mean Charlson Comorbidity Index (CCI) was 3.2 (SD ± 2.3).

**Table 1.** Demographics and Baseline Clinical Characteristics*

| **Table 1. Demographics and Baseline Clinical Characteristics\*** |  |
| --- | --- |
| **Characteristic** | **Patients (N=20)** |
| Age, mean (range) – yr | 61 (23-87) |
| Sex – no. (%) |  |
| Female | 12 (60) |
| Male | 8 (40) |
| Race – no. (%) |  |
| White | 18 (90) |
| Black | 1 (5) |
| Declined to state | 1 (5) |
| Body-mass index (BMI), mean – kg/m<sup>2</sup> | 30.1 |
| Admission Location – no. (%) |  |
| Home | 17 (85) |
| Group Home | 3 (15) |
| Insurance Type |  |
| Medicare + Medicaid | 4 (20) |
| Commercial | 5 (25) |
| Commercial + Medicare | 6 (30) |
| Medicaid | 4 (20) |
| Self-pay | 1 (5) |
| Comorbid conditions – no. (%) |  |
| Obesity | 10 (50) |
| Class I obesity (BMI: 30-34.9 kg/m<sup>2</sup>) | 6/10 (60) |
| Class II obesity (BMI: 35-39.9 kg/m<sup>2</sup>) | 1/10 (10) |
| Class III obesity (BMI: ≥40 kg/m<sup>2</sup>) | 3/10 (30) |
| Cancer | 2 (10) |
| Cardiovascular Disease | 4 (20) |
| CAD | 2 (10) |
| Atrial Fibrillation | 3 (15) |
| Congestive Heart Failure | 1 (5) |
| Chronic Liver Disease | 2 (10) |
| Chronic Kidney Disease | 4 (20) |
| Current or former smoker | 6 (30) |
| Diabetes Mellitus | 2 (10) |
| History of CVA | 1 (5) |
| Hypertension | 12 (60) |
| Pulmonary Disease | 4 (20) |
| COPD | 2 (10) |
| Asthma | 2 (10) |
| Obstructive Sleep Apnea | 4 (20) |
| Mean Charlson Comorbidity Index – score (SD) | 3.2 (2.3) |
| Mean duration of symptoms before admission – days (SD) | 6.4 (6.22) |
| Median duration of symptoms before admission – days (range) | 5 (<1-25) |
| Travel and contact history – no. (%) |  |
| Travel to endemic country in the past month | 1 (5) |
| Known COVID-19 contact(s) | 14 (70) |
| Respiratory symptoms – no. (%) |  |
| Cough | 11 (55) |
| Shortness of Breath | 11 (55) |
| Sputum Production | 3 (15) |
| Other Respiratory Symptoms | 4 (20) |
| Other symptoms – no. (%) |  |
| Fever | 10 (50) |
| Fatigue | 4 (20) |
| Rigors | 4 (20) |
| Nausea/Vomiting | 4 (20) |
| Weakness | 3 (15) |
| Decreased Appetite | 3 (15) |
| Malaise | 3 (15) |
| Abdominal Pain | 2 (10) |
| Altered Mental Status | 2 (10) |
| Diaphoresis | 2 (10) |
| Chest Pain/ Pressure | 2 (10) |
| Myalgias/Arthralgias | 2 (10) |
| Lethargy | 1 (5) |
| Dizziness | 1 (5) |
| Syncope | 1 (5) |
| Ageusia | 1 (5) |
| Headache | 1 (5) |
| Dry Mouth | 1 (5) |
| Anorexia | 1 (5) |
| Admission vital signs |  |
| Temperature $\geq 100.4$ F – no. (%) | 6 (30) |
| Heart rate $\geq 100$ BPM – no. (%) | 3 (15) |
| Respiratory rate $\geq 20$ breaths/min – no. (%) | 12 (60) |
| Oxygen saturation $\leq 93\%$ – no. (%) | 9 (45) |
| Receiving supplemental oxygen when oxygen saturation first measured | 7 (35) |

\*Plus-minus values are ± SD. IQR denotes interquartile range. ICU denotes intensive care unit.
BMI denotes body-mass index.

The mean duration of symptoms before admission was 6.4 days (SD ± 6.22, range <1-25) and one patient had a history of travel to an endemic country with the month prior to admission. Fourteen patients (70%) had contact with a known COVID-19 positive patient prior to admission. The most common symptoms upon presentation included cough (55%), shortness of breath (55%), and/or fever (subjective or objective, 50%). Additional symptoms upon presentation are listed in Table 1.

On admission, six (30%) patients had a measured temperature of ≥ 100.4^⍰^F, three (15%) had a heart rate ≥ 100 beats per minute, 12 (60%) had a respiratory rate ≥ 20 breaths per minute, and nine (45%) had an oxygen saturation ≤ 93%. Seven (35%) patients were receiving supplemental oxygen when the oxygen saturation level was measured on admission.

### Radiologic and laboratory findings

The radiologic and notable laboratory findings of these patients are described in Table 2. Fifteen (93%) patients displayed radiographic evidence of COVID-19 pneumonia on chest x-ray (CXR) or CT. Of these, four (27%) patients had diffuse, severe bilateral alveolar infiltrates on CXR.

**Table 2.**
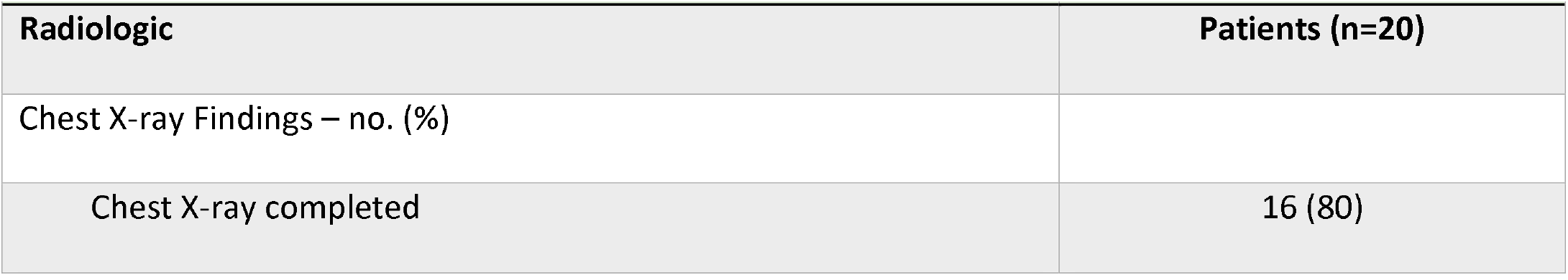
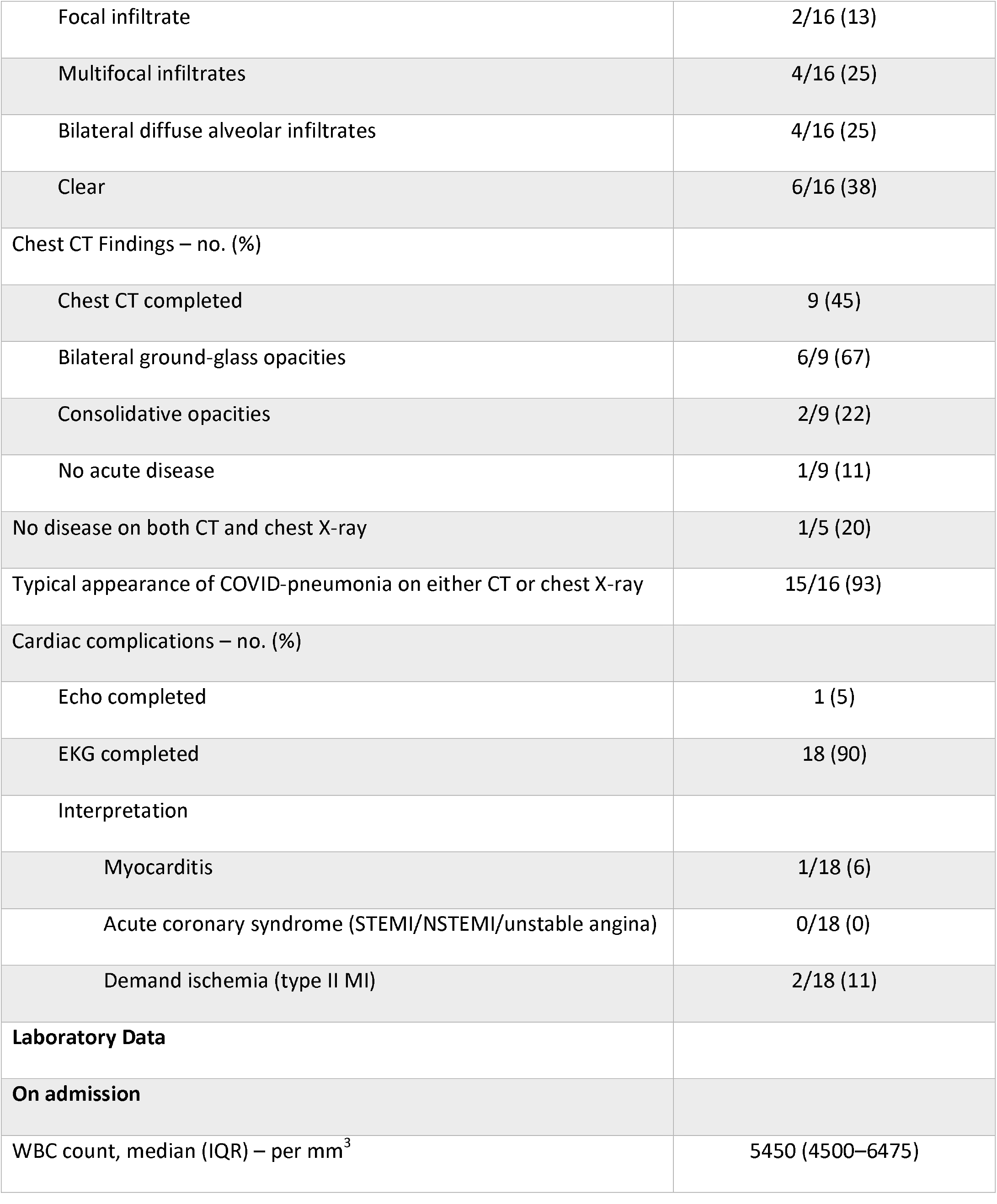
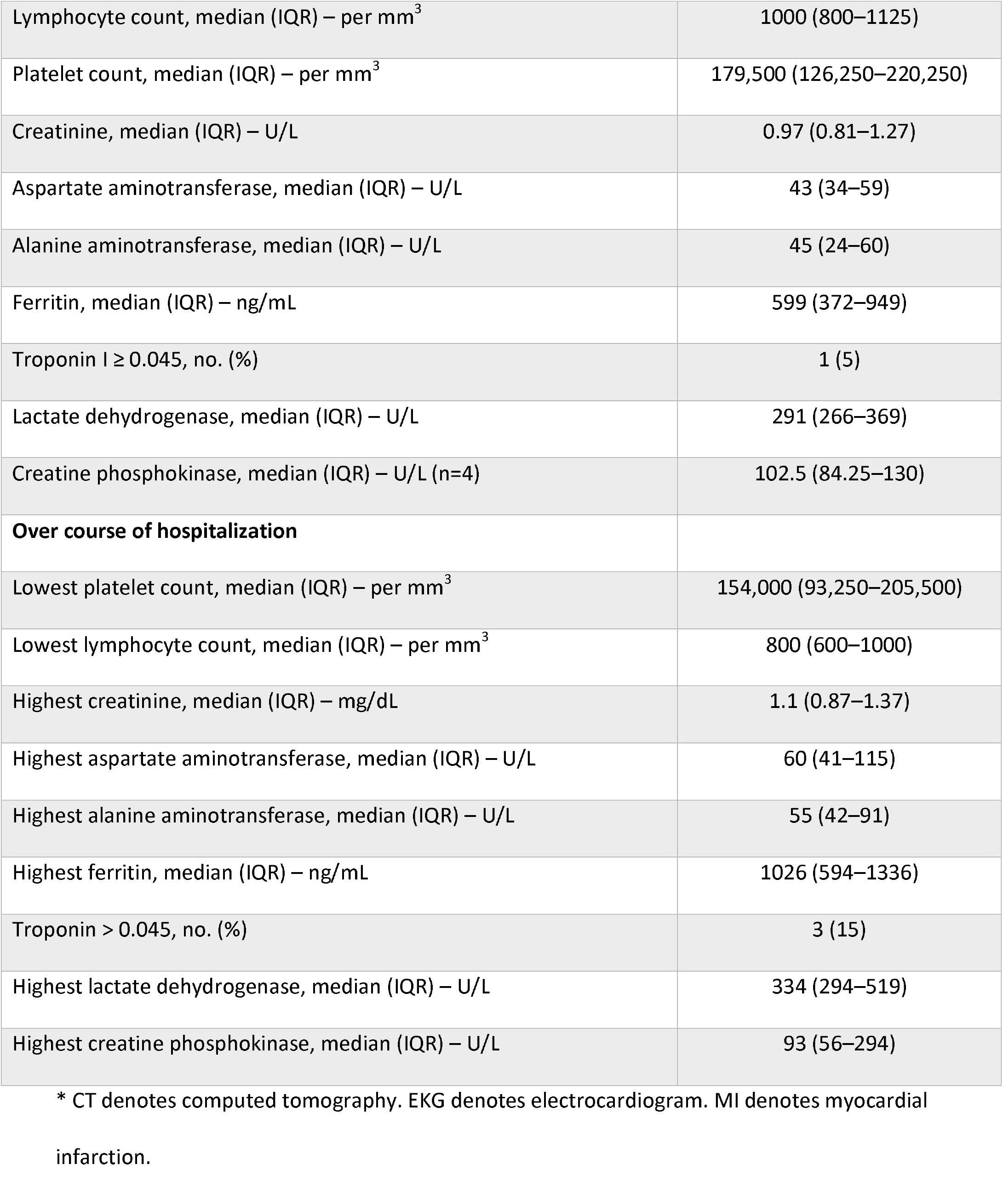
Radiologic and Baseline Laboratory Findings*

BMI denotes body-mass index.
| Table 2. Radiologic and Baseline Laboratory Findings* |  |
| --- | --- |
| Radiologic | Patients (n=20) |
| Chest X-ray Findings – no. (%) |  |
| Chest X-ray completed | 16 (80) |
| Focal infiltrate | 2/16 (13) |
| Multifocal infiltrates | 4/16 (25) |
| Bilateral diffuse alveolar infiltrates | 4/16 (25) |
| Clear | 6/16 (38) |
| Chest CT Findings – no. (%) |  |
| Chest CT completed | 9 (45) |
| Bilateral ground-glass opacities | 6/9 (67) |
| Consolidative opacities | 2/9 (22) |
| No acute disease | 1/9 (11) |
| No disease on both CT and chest X-ray | 1/5 (20) |
| Typical appearance of COVID-pneumonia on either CT or chest X-ray | 15/16 (93) |
| Cardiac complications – no. (%) |  |
| Echo completed | 1 (5) |
| EKG completed | 18 (90) |
| Interpretation |  |
| Myocarditis | 1/18 (6) |
| Acute coronary syndrome (STEMI/NSTEMI/unstable angina) | 0/18 (0) |
| Demand ischemia (type II MI) | 2/18 (11) |
| **Laboratory Data** |  |
| **On admission** |  |
| WBC count, median (IQR) – per mm<sup>3</sup> | 5450 (4500–6475) |
| Lymphocyte count, median (IQR) – per mm<sup>3</sup> | 1000 (800–1125) |
| Platelet count, median (IQR) – per mm<sup>3</sup> | 179,500 (126,250–220,250) |
| Creatinine, median (IQR) – U/L | 0.97 (0.81–1.27) |
| Aspartate aminotransferase, median (IQR) – U/L | 43 (34–59) |
| Alanine aminotransferase, median (IQR) – U/L | 45 (24–60) |
| Ferritin, median (IQR) – ng/mL | 599 (372–949) |
| Troponin I ≥ 0.045, no. (%) | 1 (5) |
| Lactate dehydrogenase, median (IQR) – U/L | 291 (266–369) |
| Creatine phosphokinase, median (IQR) – U/L (n=4) | 102.5 (84.25–130) |
| **Over course of hospitalization** |  |
| Lowest platelet count, median (IQR) – per mm<sup>3</sup> | 154,000 (93,250–205,500) |
| Lowest lymphocyte count, median (IQR) – per mm<sup>3</sup> | 800 (600–1000) |
| Highest creatinine, median (IQR) – mg/dL | 1.1 (0.87–1.37) |
| Highest aspartate aminotransferase, median (IQR) – U/L | 60 (41–115) |
| Highest alanine aminotransferase, median (IQR) – U/L | 55 (42–91) |
| Highest ferritin, median (IQR) – ng/mL | 1026 (594–1336) |
| Troponin > 0.045, no. (%) | 3 (15) |
| Highest lactate dehydrogenase, median (IQR) – U/L | 334 (294–519) |
| Highest creatine phosphokinase, median (IQR) – U/L | 93 (56–294) |

\* CT denotes computed tomography. EKG denotes electrocardiogram. MI denotes myocardial infarction.

On admission, ferritin and lactate dehydrogenase (LDH) median values were elevated at 599 ng/ml (IQR 372-949, reference range 11-264) and 291 U/L (IQR 266-369, reference range 84-246), respectively. The median lymphocyte count was at the lower limit of normal range at 1000/mm^3^ (IQR 800-1125, reference range 1000-3100). Over the course of hospitalization, the median absolute lymphocyte count decreased (800/mm^3^, IQR 600-1000) and the median ferritin (1026 ng/mL, IQR 594-1336) and LDH levels (334 U/L, IQR 294-519) increased.

Troponin was elevated in one patient (5%) on admission and three patients (15%) over the course of hospitalization. Additional admission and hospitalization laboratory findings are included in Table 2.

### Clinical course, disease severity, and treatment course

The clinical course, disease severity, and treatment course of these patients is described in Table 3. The median length of stay was six days (IQR 2-23). Four patients (20%) required ICU level care with two patients requiring ICU care for one day and with two patients requiring ICU care for 19 and 27 days, respectively.

**Table 3.** Clinical Course and Disease Severity*

| **Table 3. Clinical Course and Disease Severity\*** |  |
| --- | --- |
| **Length of care** | **Patients (n=20)** |
| Median length of hospitalization – days (IQR) | 6 (2-23) |
| ICU-level care required – no. (%) | 4 (20) |
| Mechanically ventilated – no. (%) | 2 (10) |
| **Patient 1** |  |
| Length of ICU care – days | 19 |
| Length of invasive mechanical ventilation – days | 15 |
| **Patient 2** |  |
| Length of ICU care – days | 27 |
| Length of invasive mechanical ventilation – days | 24 |
| **Patient 3 and 4** |  |
| Length of ICU care – days | 1 |
| **Severity of illness -- NIH criteria – no. (%)** |  |
| Non-severe | 4 (20) |
| Severe | 9 (45) |
| Critical | 7 (35) |
| **Critical criteria qualification:** |  |
| Respiratory Failure | 7/7 (100) |
| Septic Shock | 3/7 (43) |
| Multiple Organ Dysfunction | 5/7 (71) |
| Acute kidney injury | 6/7 (86) |
| Transaminitis | 16 (80) |
| **Supplemental oxygen requirements – no. (%)** |  |
| No supplemental oxygen requirement | 7 (35) |
| Low flow oxygen (nasal cannula) | 5 (25) |
| High flow oxygen (nasal cannula) | 3 (15) |
| Non-invasive ventilation (CPAP/BIPAP) | 3 (15) |
| Invasive Mechanical Ventilation | 2 (10) |
| Proned | 0 |
| **Treatments received** |  |
| **COVID-19 Targeted Treatments** |  |
| Hydroxychloroquine – no. (%) | 5 (25) |
| Azithromycin – no. (%) | 2 (10) |
| Steroids – no. (%) | 4 (20) |
| Mean length of steroid therapy – days | 8.8±5.6 |
| Tocilizumab – no. (%) | 9 (45) |
| Mean number of Tocilizumab doses | 2±0.7 |
| Convalescent Plasma – no. (%) | 3 (15) |
| **Supportive Care Medications** |  |
| Vasopressors – no. (%) | 3 (15) |
| Inhaled bronchodilators – no. (%) | 5 (25) |
| **NIH Ordinal Scale – score** |  |
| Mean on admission | 4.3±0.8 |
| Mean lowest during hospitalization | 3.8±1.2 |

\*Plus-minus values are ±SD. IQR denotes interquartile range. ICU denotes intensive care unit.
CPAP denotes continuous airway pressure. BIPAP denotes bilevel positive airway pressure.

Using the NIH COVID-19 severity of illness categories^11^, nine (45%) patients met NIH criteria for severe illness and seven (35%) met criteria for critical illness. Of the seven patients with critical illness, all (100%) met criteria for respiratory failure, three (43%) developed septic shock, five (71%) developed multi-organ dysfunction, and six patients (86%) developed acute kidney injury^10^. 16 (80%) patients developed transaminitis.

Cardiac complications included demand ischemia in two (10%) patients and myocarditis in one (5%) patient. No patients developed acute coronary syndrome and there were no other thromboembolic complications.

Seven (35%) patients did not require supplemental oxygen, five (25%) received low flow supplemental oxygen, three (15%) received high flow oxygen via high flow nasal cannula, three (15%) received non-invasive mechanical ventilation, and two (10%) received invasive mechanical ventilation. No patients were proned.

Five patients (25%) received hydroxychloroquine, two (10%) received azithromycin (in addition to hydroxychloroquine), four (20%) received systemic corticosteroids, nine (45%) received tocilizumab, and three (15%) received convalescent plasma. Of the patients who received tocilizumab, the mean number of doses per patient was two (SD ±0.74). Five patients (25%) received inhaled bronchodilators and three (15%) received vasopressors.

The mean NIH ordinal scale score was 4.3 on admission (SD ±0.78), with the mean lowest score over the course of hospitalization being 3.8 (SD ±1.2). One (5%) patient met criteria for invasive mechanical ventilation, but this was not consistent with their goals of care. The patient died after transitioning to comfort measures. The remaining 19 (95%) patients survived and no patients were transferred to other hospitals.

## DISCUSSION

### Clinical Approach and Challenges

This regional, single center study represents to our knowledge the first template for delivering care to hospitalized COVID-19 patients in a rural area. Our approach included the creation of a regional hub and spoke system for routing patients to the flagship hospital in our health system as well as creation of a multidisciplinary team with experience in hospitalist medicine, rheumatology, infectious diseases and inpatient pharmacy. This allowed us to centralize a number of limited resources (clinical expertise, medications), to stay abreast of developing research on SARS-CoV-2 infection and to quickly adapt to changing guidelines for the treatment of COVID-19. Fortunately, although there was a high rate of comorbidities in this case series, the death rate was 5%, no patients were transferred to outside hospitals and no full code patients died. This was in the setting of lacking access to remdesivir, dialysis services and extracorporeal membrane oxygenation (ECMO) in our health system.

One of our team’s key challenges was in adapting to a quickly changing standard of care. For example, five patients initially received hydroxychloroquine and/or azithromycin, but upon publication of a retrospective VA study that suggested no benefit to this approach^12^ we stopped using both of these medications. In light of an encouraging case series from China^13^ and encouraging data from an open label randomized controlled tocilizumab trial in France^14^, we also used tocilizumab for nine of our patients. However, the high cost of this medication, coupled with both our health system’s dropping revenues in the early part of the COVID-19 pandemic and the uncertainty of third party reimbursement, made the decision of when to administer tocilizumab particularly challenging. In response to this, our team worked closely with the SLHS’s clinical research department to develop clinical trial opportunities for our hospitalized COVID-19 patients, and this case series concludes at the starting point of our health system’s first COVID-19 inpatient trial. This underscores the need for and importance of clinical research departments in rural health systems. Had we been part of a larger COVID-19 clinical trial earlier in the pandemic, the question of cost would not have been part of our clinical decision-making process.

### Broader Applicability

There are several lessons to be learned from our rural inpatient COVID-19 experience. The first is that small teams of providers can be quickly assembled in response to unexpected medical emergencies and events. This is relevant to rural areas in the U.S., the majority of which have local health systems that lack the resources of academic centers^15^. The nature of these small teams is also important. In particular, physicians who are generalists within their fields can be used to meet unexpected needs. In our team’s case, a rheumatologist with experience in the management of interstitial lung disease and two hospitalists with experience in critical care medicine all served broad roles within their respective fields. These are just several examples, and what is even more important than our team’s COVID-19 experience is the larger lesson that in the setting of resource limitation small teams of generalists can be used to respond to a wide range of medical events.

Another lesson is that having the infrastructure and experience to conduct clinical trials is invaluable in a pandemic. At a time when the majority of rural sites lack this infrastructure^6-7^ there is often a divergence between national treatment guideline^8-9^ and the on the ground reality for rural hospitals. In our own case, outside of convalescent plasma we were not enrolled in any clinical trials over the dates of this case series. We were unable to establish access to remdesivir for any of our patients and IV tocilizumab was obtained only after a significant procurement effort by our inpatient COVID-19 team. Furthermore, there was on average a nearly two week delay between the day of request and the day of receipt of convalescent plasma for the patients in this series. Fortunately, we were ultimately able to develop a clinical trial opportunity for our hospitalized COVID-19 patients.

A third and overarching lesson is that crisis management either in or out of a pandemic requires adequate networks of care. Such networks must include systems for centralizing and streamlining healthcare resources, good communication between physicians and advanced practice providers at hub and spoke hospitals, clinical research infrastructure and expertise, and ideally strong relationships between rural and regional academic physicians.

### Limitations

Our case series has multiple important limitations. These include our small sample size, the lack of a control group, the retrospective nature of this series and the relatively small numbers of patients who received immunomodulatory medications. The chief goal of this report, however, is not to shine a light on any one treatment strategy for COVID-19 patients but rather to give an example of what a successful prototype for a rural pandemic response system might entail.

It should also be noted that our rural COVID-19 experience has been atypical. Whereas in our health system we had the subspecialty expertise and access to medications to at least approximate clinical trial-like conditions for our patients, the majority of rural health systems lack such resources. And yet, our health system serves one of the most vulnerable populations to COVID-19 in New York state^16^ and the patients in this series provide at least a reasonable (if small) representation of the rural U.S. population^17^. As this is to our knowledge only the second reported case series of hospitalized COVID-19 patients in a rural area^18^, we believe that our experience can help provide a template through which to structure future crisis management approaches in rural areas. Furthermore, while this case series may not reflect the typical resources of a rural hospital, we also believe that such resources can be developed. In our health system’s case, we have been able to create a clinical research department and recruit the subspecialists on our COVID-19 inpatient management team within the past five years.

### Recommendations and Conclusions

A potential rural pandemic response system of the future might involve a hub and spoke system in which a larger rural hospital (the hub) would provide the resources to treat the sicker COVID-19 patients in its region. In turn, smaller rural hospitals (the spokes) within that region could refer sicker COVID-19 patients to the hub hospital and consult with a multidisciplinary team at the hub hospital on patients who could be managed locally (Figure 1). This has been the model for COVID-19 care delivery in our region, and our own experience suggests that, with the right impetus, the resources for such a model can be developed. An improved version of this model would also include a backup COVID-19 consultation team at a regional academic center, the incorporation of patient engagement and machine learning tools into care delivery, and research alliances that connect hub and spoke hospitals with larger academic partners (Figure 1).

**Figure 1.**
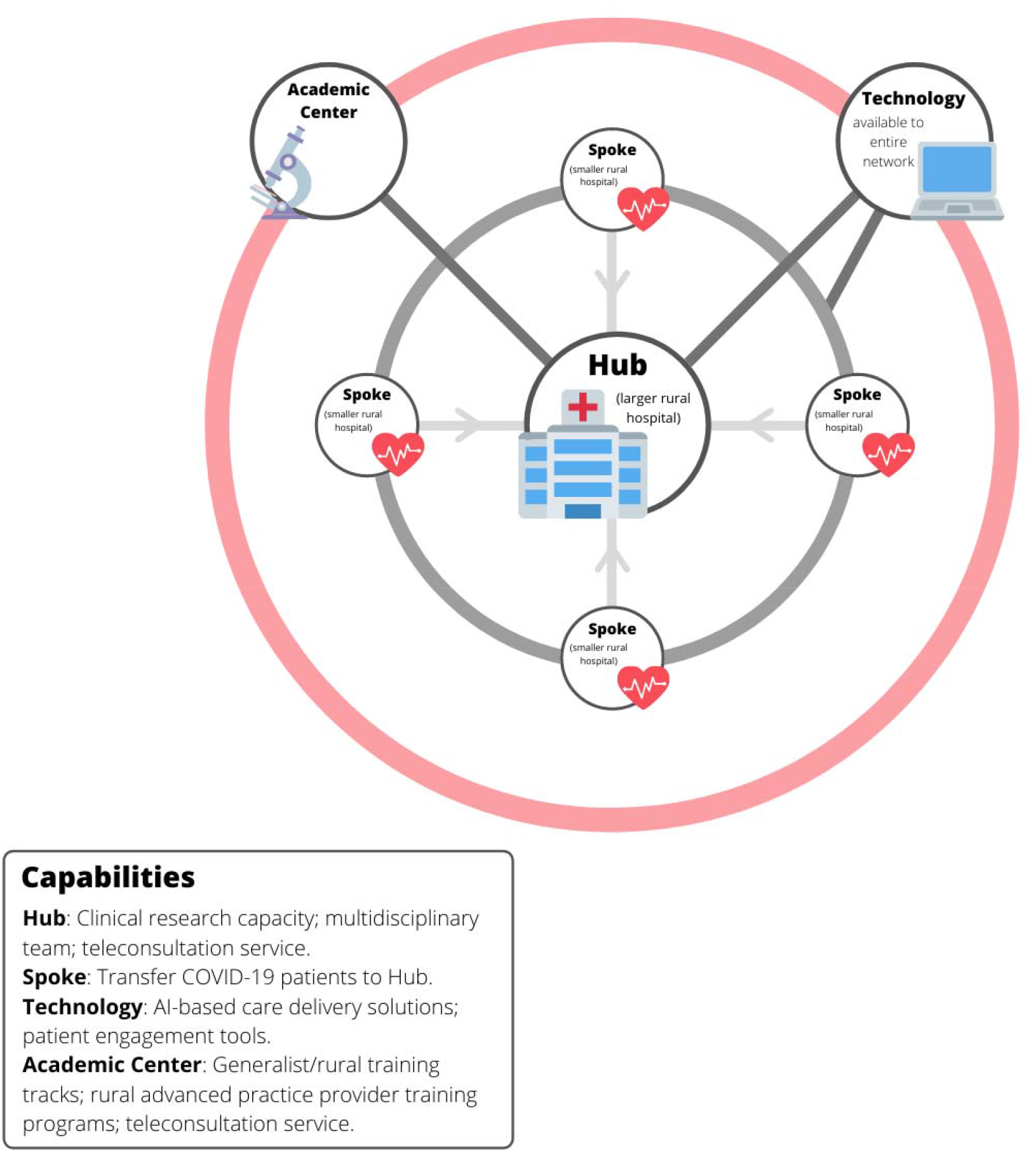
Hub and Spoke model for rural care delivery

Our experience also suggests that the clinical approach of the physicians on a rural pandemic response team matters. A small team of generalist physicians can in a crisis provide a level of care comparable to that found in academic centers. As the rural subspecialty gap is likely to remain severe^19-21^, efforts to develop both specialists and subspecialists with a generalist approach within their respective fields will be essential.

The COVID-19 pandemic has given fresh exposure to a number of longstanding health and health care disparities in the U.S^22-23^.. One of these health care disparities is that of rural Americans and the often under-resourced health systems that serve them. Based upon our experience during the initial phase of COVID-19 in our area, we would advise rural health policymakers and our colleagues in academia to consider the development of regional hub and spoke models for rural settings, to invest in specialty and subspecialty training that is targeted to rural areas, to invest in regional and national clinical rural research networks and to continue to foster strong working relationships between rural physicians and regional academic centers. The development of strong clinical and research networks will help equip rural areas to respond not only to current and future pandemics, but more broadly to any new care delivery challenges that they face. We hope that our experience can help guide further conversation and the development of further guidelines for treating both COVID-19 and other pandemics and events in rural areas in the future.

## Data Availability

The authors declare that all data supporting the findings of this study are available within this article.

